# Wearable-derived daily steps and first recorded cardiovascular diagnoses: outpatient and acute care in All of Us

**DOI:** 10.64898/2026.10.29.26366804

**Authors:** Kathy Xu, Annie Qu

## Abstract

Wearable-derived daily step counts are associated with cardiovascular health, but associations with the setting of cardiovascular disease (CVD) recording are less clear. We conducted a retrospective landmark cohort study using linked Fitbit, survey, and electronic health record data from the All of Us Research Program. Steps were averaged over the year before baseline. Eligibility required post-baseline sleep coverage and survey completeness, and classifiable pre-baseline smoking and drinking information; former smokers were excluded. After excluding participants with primary or secondary CVD recorded before baseline and 1,470 with either status during the first post-baseline year, outcomes were classified by the first qualifying primary CVD record during years two and three; a single qualifying outpatient record was sufficient. Among 8,292 participants, 5,780 had no recorded CVD, 2,127 had outpatient recording, and 385 had acute-care recording. Multinomial models adjusted for age, sex at birth, smoking, and drinking categories. Each one-standard-deviation increase in step count (3,299 steps/day) was associated with lower odds of outpatient recording versus no recorded CVD (odds ratio 0.786, 95% confidence interval 0.744–0.830), acute-care recording versus no recorded CVD (0.659, 0.586–0.742), and acute-care versus outpatient recording (0.839, 0.742–0.948). Both associations relative to no recorded CVD remained inverse across the tested hypothetical allocations of 455 missing-setting cases, whereas the acute-care-versus-outpatient point estimate approached or crossed the null under some scenarios. Secondary exploratory mediation analyses using a pooled score combining sleep, mental-health, and clinical-burden measures identified indirect associations for both settings versus no recorded CVD, but the between-setting indirect contrast was inconclusive. In this selected cohort, the findings highlight the importance of endpoint definition and recording completeness when interpreting wearable–EHR associations.

**Author summary:** We asked whether daily steps were related to where cardiovascular disease was subsequently recorded as a primary diagnosis: in outpatient care or in a hospital or emergency department. We linked Fitbit, survey, and electronic health record data from the All of Us Research Program. Our selected cohort met wearable and survey data requirements and had classifiable smoking and drinking information; former smokers were excluded. Participants had no cardiovascular diagnoses recorded before baseline or during the following year. We averaged steps over the year before baseline and assessed recording settings during years two and three. Higher step counts were associated with lower odds of both recording settings relative to no recorded cardiovascular disease. These associations remained inverse across the tested hypothetical allocations of cases with missing settings, but the estimated difference between settings weakened or changed direction under some allocations. A secondary exploratory analysis found statistical indirect associations involving a health-burden score, but the indirect comparison between settings remained inconclusive. The findings apply to this selected cohort, rather than all wearable users, and show why recording context and completeness matter when interpreting wearable–health-record associations. Care settings reflect documentation and care use, not simply disease severity.

## Introduction

Cardiovascular disease (CVD) remains a major cause of morbidity and mortality. Wearable devices capture habitual physical activity outside clinical encounters, and prospective studies have associated higher objectively measured activity and daily step counts with lower cardiovascular risk [1–3]. In the All of Us Research Program, longitudinal Fitbit-derived step counts linked with electronic health records (EHRs) have also been associated with chronic disease risk [4]. Less is known about whether these associations differ by the setting of the first recorded primary CVD diagnosis: outpatient care or a hospital or emergency department. Recording setting is not a direct measure of severity: it can also reflect disease subtype, access to care, care-seeking, and documentation.

Unlike prior studies that examined aggregate cardiovascular incidence, this study asks whether wearable-derived activity is differentially associated with where CVD is first recorded in the EHR, while explicitly evaluating the sensitivity of this setting contrast to incomplete encounter-setting information. Cases without a classified setting cannot contribute directly to an outpatient-versus-acute-care comparison, and their exclusion may affect the estimated association. We therefore paired the known-setting analysis with hypothetical allocations of missing-setting cases to examine which associations retained their direction and which depended on the allocation assumptions. Research on activity, sleep, mental health, and cardiovascular outcomes also motivates considering related health domains jointly [5–8]. Studies of joint activity–sleep associations and sleep–depression–CVD associations provide further context [9, 10]. As a complementary analysis, we used one pooled principal component analysis (PCA) score from sleep, mental-health, and clinical-burden domains to explore model-based indirect associations. Sex and continuous-age modification were secondary questions.

## Materials and methods

### Study design and participants

We conducted a retrospective *t*_1_-landmark cohort study using linked electronic health record (EHR), survey, and Fitbit data from participants in the *All of Us* Research Program, a nationwide research cohort in the United States. The study used the *All of Us* Registered Tier Curated Data Repository version 8, with a data cutoff of Oct 1, 2023. Data were accessed for this study through the Researcher Workbench from 28 November 2025 to 19 June 2026. During upstream processing, daily wear time was estimated by counting distinct minutes with recorded heart-rate data and dividing by 60; these minutes did not have to be consecutive. A Fitbit day was considered valid when it contained at least 600 such minutes (10 hours of estimated wear) and at least 100 recorded steps, following the valid-day thresholds used in previous All of Us Fitbit research [4]. We defined a valid calendar month as one containing at least 21 valid days; this monthly threshold and the requirement for at least nine valid months were study-specific criteria applied upstream during cohort preparation. Together, these criteria required at least three weeks of valid observations per contributing month and coverage of at least three quarters of the exposure year, while allowing intermittent non-recording. For each participant, candidate baseline dates were the first days of successive calendar months, evaluated in chronological order. We selected the earliest candidate date as *t*_0_ when both Fitbit requirements were met: at least 12 months of prior observation and at least nine valid months among the 12 complete calendar months immediately preceding that date. The nine valid months did not have to be consecutive; eligibility was assessed across the full rolling 12-month window, not from a single qualifying month. Accordingly, a participant *i* was eligible only if both conditions were met:

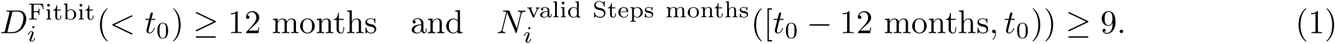

This common rule was applied to participants who subsequently had CVD recorded and to participants without recorded CVD. Baseline dates (*t*_0_) in the analytic cohort ranged from February 1, 2018, to October 1, 2020. Observation duration alone was insufficient for inclusion: participants with at least 12 months of pre-*t*_0_ Fitbit observation were excluded if they had fewer than nine valid Steps months within the designated 12-month exposure window. Eligibility also required EHR availability through the common endpoint at *t*_0_ + 36 months, operationalised as a last available EHR record on or after that endpoint. No participant in the extracted analytic cohort had a recorded death. Prior CVD was assessed using all available EHR history before *t*_0_, from each participant’s earliest available EHR record up to, but not including, *t*_0_. Participants with a primary or secondary CVD diagnosis recorded in this interval were excluded upstream. For cohort screening, recorded CVD refers to these two diagnosis statuses, as detailed below. This criterion establishes absence of such records within the available history, rather than lifetime absence of disease.

The participant-specific baseline, *t*_0_, marked the end of the 12-month Fitbit exposure-assessment window. The health-burden ascertainment cutoff was *t_M_*= *t*_0_ + 1 month, and the outcome landmark was *t*_1_ = *t*_0_ + 12 months. All outcome groups used these same windows. Only pre-*t*_0_ Fitbit records contributed to Steps; health-burden ascertainment used records during [*t*_0_*, t_M_*), followed by an event-free lag until *t*_1_. Neither diagnoses after *t*_0_ nor outcomes contributed to the exposure definition. The prespecified sequence was:

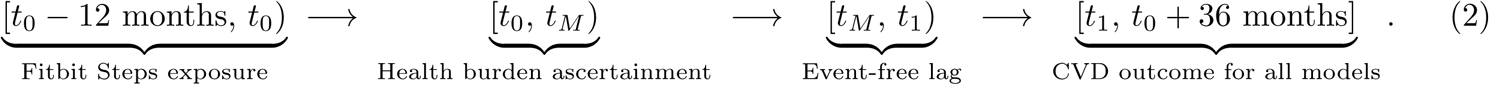

The available cohort counts begin with 10,365 participants satisfying the *t*_0_ Fitbit and EHR criteria after exclusion of prior recorded CVD. This reported starting cohort already incorporated the diagnosis-status exclusions described below, complete age and sex-at-birth information, at least seven valid main-sleep days during [*t*_0_*, t_M_*), complete depression and anxiety survey responses, eligible recorded smoking status, and non-missing, determinate pre-baseline drinking-frequency information. Participants classified as former smokers or without any eligible pre-baseline smoking-status record, and those with missing or indeterminate drinking-frequency information, were excluded upstream, before the counted flow began. During cohort preparation within the Researcher Workbench, 1,470 participants with primary or secondary CVD recorded during [*t*_0_*, t*_1_) were removed. The prepared cohort comprised 8,895 participants without recorded CVD before *t*_1_, and the common outcome risk period for M1, the Steps-by-sex model, M2, and all exploratory mediation analyses was [*t*_1_*, t*_0_ + 36 months]. M1 estimated the adjusted Steps–outcome association without the health-burden score, whereas M2 included the health-burden score in the same *t*_1_-landmark cohort. Diagnosis-status screening is described below, and a consolidated sequence of cohort-construction rules is provided in S1 Text.

The prepared cohort included 5,780 participants with neither primary nor secondary CVD recorded through *t*_0_ + 36 months (the no-recorded-CVD category) and 3,115 participants with a first qualifying primary CVD record during [*t*_1_*, t*_0_ + 36 months]. The encounter setting of that record determined the outcome category: hospital or emergency-department encounters were classified as acute care, and office or outpatient encounters as outpatient care. When primary CVD records occurred in more than one setting on the same earliest date, acute care took precedence. A second outpatient code was not required. We use the concise terms acute-care recording and outpatient recording for these primary-diagnosis outcome categories. Among the 3,115 cases, 385 had an acute-care recording and 2,127 had an outpatient recording; 148 had another or unknown setting, and 455 had missing setting information. Excluding these 603 unclassified cases yielded 8,292 participants for the primary multinomial analysis (Fig 1). Study size was determined by the available records meeting these eligibility and outcome-classification criteria.

**Fig 1.**
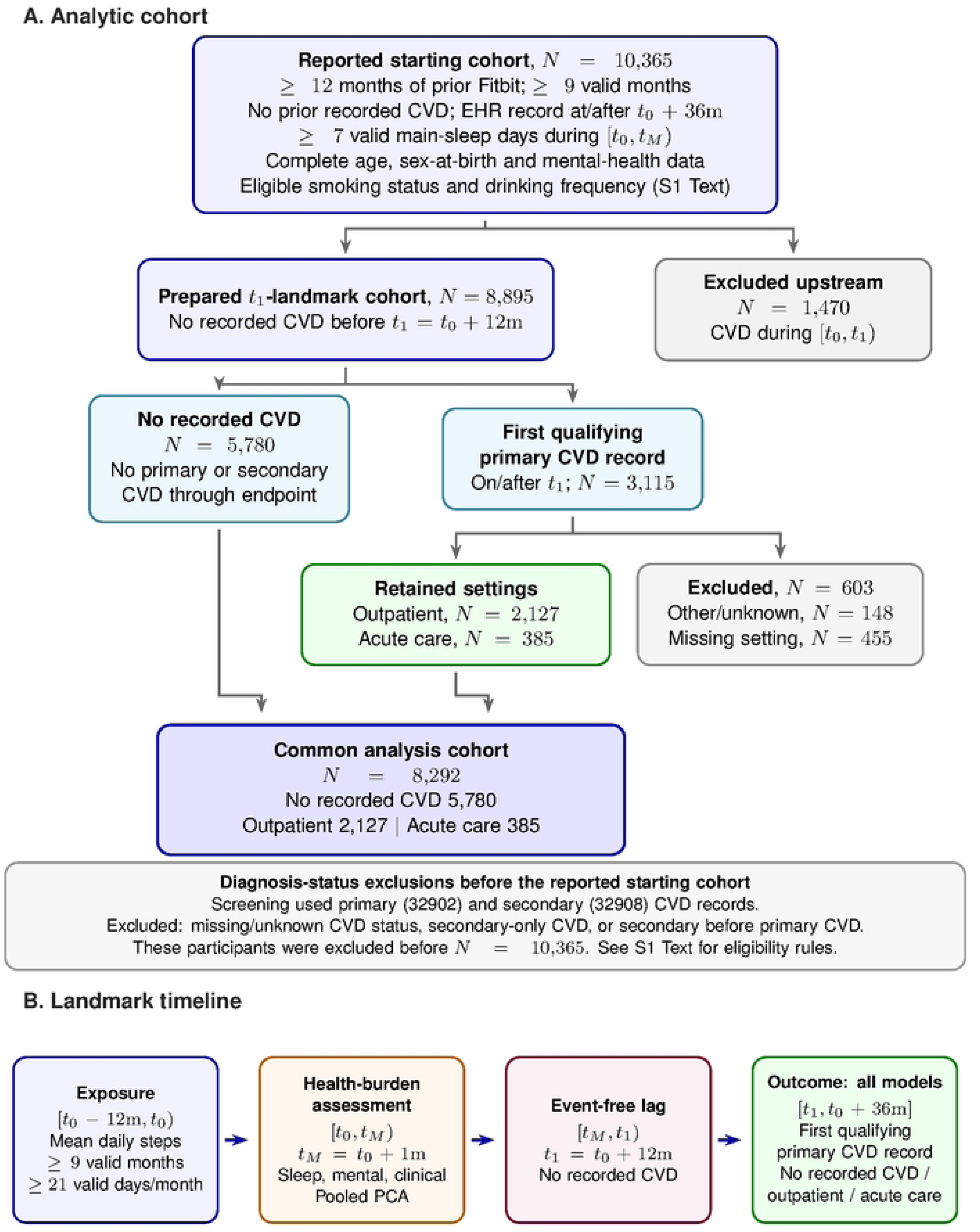
Study cohort flow and landmark design. (A) The reported starting cohort (*N* = 10,365) already met the Fitbit, EHR, diagnosis-status, demographic-completeness, sleep-coverage, survey-completeness, smoking-status, and drinking-frequency eligibility criteria, with no prior primary or secondary CVD recorded. After 1,470 first-year CVD exclusions and 603 unclassified-setting exclusions, 8,292 participants entered the primary association and exploratory mediation models. Earlier eligibility exclusions are outside this reported flow; screening rules are described in Methods and S1 Text. Cases were classified by their first qualifying primary CVD record, and participants without recorded CVD had neither primary nor secondary CVD through the endpoint. (B) Steps were assessed during [*t*_0_ − 12m*, t*_0_) and health burden during [*t*_0_*, t_M_*), with *t_M_* = *t*_0_ + 1 month. Following the event-free lag, all models assessed outcomes during [*t*_1_*, t*_0_ + 36m], where *t*_1_ = *t*_0_ + 12 months. CVD=cardiovascular disease; EHR=electronic health record.

### Ethics and data governance

This study was a secondary analysis of existing, deidentified data from the *All of Us* Research Program Registered Tier Curated Data Repository, version 8. The *All of Us* Research Program’s protocol and informed-consent procedures were reviewed and approved by its Institutional Review Board, and participants provided electronically documented informed consent. The present study involved no participant recruitment, direct interaction, intervention, or new collection of data or biospecimens.

We accessed the data through the secure *All of Us* Researcher Workbench under the institutional Data Use and Registration Agreement between Vanderbilt University Medical Center and the Regents of the University of California, on behalf of its Santa Barbara campus, effective 8 August 2023. We completed the required responsible-conduct-of-research training and agreed to the *All of Us* Data User Code of Conduct. We had no access to direct identifiers or the reidentification key at any time during this secondary analysis and did not attempt to identify or contact participants. Under the *All of Us* data-access framework, individual projects using deidentified Researcher Workbench data do not require separate project review by the *All of Us* Institutional Review Board.

Potential privacy risks from inadvertent identification were mitigated through deidentification, controlled data access, and reporting limited to aggregate results with small-cell and complementary suppression. The public repository contains downstream analysis code, variable definitions, and environment documentation, but no participant-level data. Access to the restricted source data requires authorisation through the Researcher Workbench.

### Outcomes and exposure

The outcome was the encounter setting of the earliest qualifying primary CVD record during [*t*_1_*, t*_0_ + 36 months]. CVD records with primary or secondary diagnosis status (condition_status_concept_id 32902 and 32908, respectively) were used for participant screening and eligibility for the no-recorded-CVD category. Among eligible cases, only primary records determined outcome date and setting. S9 Table lists the disease concepts and diagnosis-status codes, and S1 Text describes the data sources, variable derivation, and coverage of the public analysis code.

Diagnosis records were matched to eligible participants and their landmarks by person_id, with condition_start_date restricted to *t*_1_ through *t*_0_ + 36 months, inclusive. Records in condition_occurrence were linked to visit_occurrence by visit_occurrence_id and person_id using a left join, retaining diagnoses without a matched encounter. Setting was assigned from visit_concept_id; the code mapping is reported in S1 Text. The earliest qualifying diagnosis date was selected before excluding unclassified settings, with acute care taking precedence among records on that same date. A later record with a known setting did not replace an earlier qualifying record solely because the earlier setting was unclassified.

Before the reported starting cohort of 10,365 participants was formed, participant-level screening compared primary and secondary CVD records within each person_id. Participants whose earliest secondary CVD record preceded their earliest primary CVD record were excluded entirely, limiting the target recording pattern to a qualifying primary record without an earlier secondary CVD record. Participants with secondary CVD but no primary CVD were excluded because they met neither the primary-record case definition nor the no-recorded-CVD definition. Participants with missing or unknown CVD diagnosis status were also excluded. Independently of the record-order rule, participants without recorded CVD were required to have neither primary nor secondary CVD recorded from the start of available EHR history through *t*_0_ + 36 months. These diagnosis-status exclusions preceded the counted flow and were distinct from the later exclusion of 603 primary-CVD cases with unclassified encounter setting.

CVD was identified using EHR diagnosis concepts selected by cardiovascular condition names and corresponding codes. Diagnosis-code identifiers were not retained in the prepared analytic dataset. The broad phenotype encompassed coronary and ischaemic heart disease (including myocardial infarction, angina, and acute coronary syndromes); cerebrovascular disease (including stroke); peripheral arterial disease; heart failure; cardiac arrhythmias; cardiomyopathy; valvular heart disease; rheumatic heart disease; congenital heart disease; aortic disease; pericardial disease; and venous thromboembolism (deep vein thrombosis or pulmonary embolism). These disease groups could overlap, but each participant contributed one classification based on the first retained primary CVD record. Participant-level CVD subtype was not retained. Hypertension was excluded from the outcome phenotype and considered only in the post-*t*_0_ clinical-burden domain. The phenotype included the 13 defining disease concepts in S9 Table and their descendants, subject to the stated outcome exclusions.

The common outcome risk period for M1, M2, and the exploratory mediation analyses was [*t*_1_*, t*_0_ + 36 months]. With *t*_1_ = *t*_0_ + 12 months, this was a 24-month risk period within the fixed 36-month post-*t*_0_ horizon. Among participants satisfying the eligibility and diagnosis-position rules above, let *T^P^* be the date of the first qualifying primary CVD record and *S^P^* its setting. The common three-category outcome was

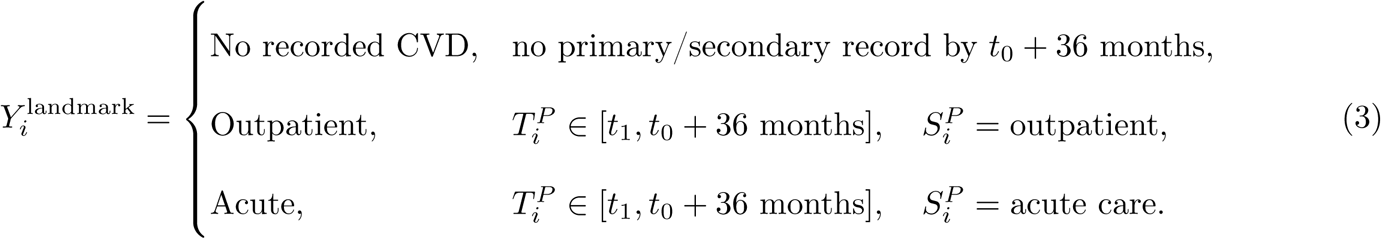

Participants with primary or secondary CVD recorded during [*t*_0_*, t*_1_) had been excluded upstream, so all analysed participants had no recorded CVD before the *t*_1_ landmark. Cases with another, unknown, or missing setting of their first qualifying primary CVD record were excluded from analyses requiring the three-category outcome.

EHR-based clinical-burden indicators were coded as 1 when a qualifying diagnosis was recorded during the specified window and 0 when none was recorded. A zero therefore denotes no recorded diagnosis, not confirmed absence of the condition. The sleep and mental-health indicators had separate device- and survey-based definitions, respectively, as described below. Requiring an EHR record on or after *t*_0_ + 36 months provided evidence of continued EHR contact but did not establish continuous capture throughout follow-up.

The primary exposure was habitual physical activity measured using Fitbit-derived daily step counts. For every participant, including participants without recorded CVD, mean daily step count was calculated from valid Fitbit observations during the common 12-month interval [*t*_0_ − 12 months*, t*_0_). Inclusion required at least nine valid calendar months, each containing at least 21 valid days, within this interval; participants contributing fewer than nine valid months were excluded. Mean daily step count was calculated across observations on valid days in the qualifying months. No diagnosis or outcome information after *t*_0_ was used to define the exposure. Mean daily step count was standardised once in the pooled complete analytic cohort as

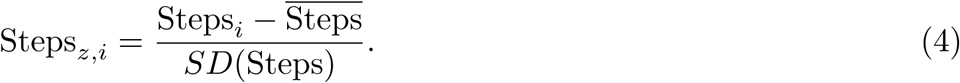

Regression ORs for Steps represent a 1-SD increase in mean daily step count, corresponding to approximately 3299 steps/day. G-computation probability contrasts specifically compare the pooled mean with 1 SD above that mean, approximately 7323 versus 10622 steps/day; they do not represent every possible 1-SD increase from an arbitrary starting value.

### Health-burden components and covariates

Device-derived sleep, self-reported mental-health diagnostic history, and EHR clinical burden were ascertained during the first post-baseline month, [*t*_0_*, t_M_*), and combined into a post-baseline health-burden score evaluated as a candidate mediator. Their different measurement sources and definitions are described below.

Sleep was a study-defined low-sleep-efficiency indicator during [*t*_0_*, t_M_*). For each main-sleep episode, efficiency was total minutes asleep (minute_asleep) divided by minutes in bed (minute_in_bed) from the same episode, using total sleep time rather than classic-specific asleep-level duration [11]. Eligibility required at least seven valid main-sleep days. Among eligible participants, Sleep was coded 1 when this ratio was ≤ 0.60 on strictly more than 75% of valid days and 0 otherwise; code 0 indicated that this threshold was not met, rather than established normal sleep. Missing or invalid days were omitted from the denominator. The indicator described recurrent low recorded efficiency, not a clinical diagnosis or the composite Fitbit Sleep Score. The complete formula, coverage rules, and operational rationale are provided in S1 Text.

The depression and anxiety indicators came from the All of Us Personal Medical History survey, whose mental-health and substance-use checklist asked about conditions previously diagnosed by a doctor or health-care provider. The relevant response options were “Depression” and “Anxiety reaction/panic disorder”; the latter is abbreviated as anxiety below. The item wording is documented in the publicly available October 12, 2018 questionnaire [12]. Participants completed the survey during [*t*_0_*, t_M_*); these were self-reported diagnostic histories, not EHR diagnoses or symptom-scale classifications. Mental burden was their sum,

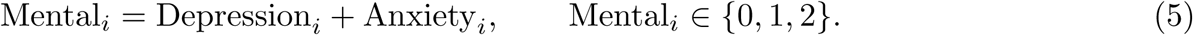

Participants with missing depression or anxiety responses were excluded upstream before creation of the analytic dataset; missing responses were not coded as absence of a diagnosis. Among eligible respondents, a value of 0 indicated neither diagnosis was reported, 1 indicated one of depression or anxiety was reported, and 2 indicated both were reported. The survey-assessment window did not imply that either diagnosis first occurred during that month or after the Steps exposure.

Clinical burden was the sum of five EHR diagnoses recorded during [*t*_0_*, t_M_*):

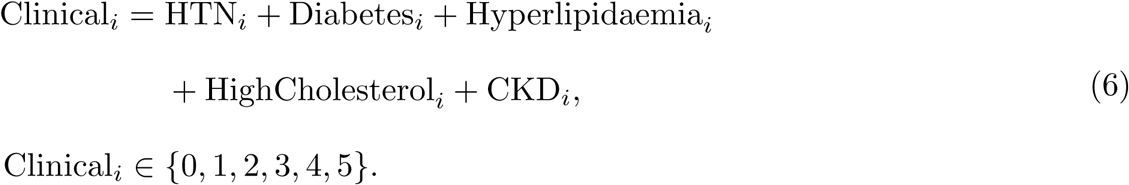

These diagnoses were components of clinical burden only when recorded during the health-burden ascertainment window; records on or after *t_M_* were not used. Component definitions are detailed in S1 Text; no primary/secondary diagnosis-status restriction was applied. Each component contributed at most one point, and components were not mutually exclusive. Hyperlipidaemia and high cholesterol used exact matches to their respective diagnosis concepts without descendant expansion; neither indicator was assigned from the other. No prior-diagnosis washout was applied, so a window-specific record could represent a longstanding condition recorded again or a first recorded diagnosis.

The adjustment set ***C****_i_* comprised age, sex at birth, recorded smoking status, and drinking categories. Covariate adjustment was constrained by the available data; measures of comorbidity and health-care utilisation preceding the Steps exposure window were unavailable in the analytic extract. Race and ethnicity were unavailable in the prepared analytic extract and were therefore neither reported descriptively nor included in covariate adjustment. Age at *t*_0_ was calculated in completed years using the birth year, month, and day extracted from birth_datetime. The sex_at_birth field contained Female and Male as its only non-missing categories; Female was the model reference. Participants with missing or NA age or sex-at-birth information were excluded upstream, before the reported starting cohort. Smoking status and drinking categories used the most recent eligible information recorded before *t*_0_; no information recorded during the health-burden or outcome windows was used for covariate adjustment. Smoking was derived from EHR observation records with observation_concept_id=4275495, using the status in value_as_concept_id. Never-smoker and non-smoker concepts were coded 0, and moderate, heavy, daily, occasional, and unspecified smoker concepts were coded 1. The reference category was not restricted to lifetime never smokers; unspecified smoker status did not establish detailed current smoking behaviour [13]. The binary scheme did not include a separate former-smoker category; explicitly recorded former smokers were excluded rather than combined with either coded group. Concept mappings and upstream status exclusions are detailed in S1 Text. Alcohol-use frequency was obtained from the most recent eligible pre-*t*_0_ All of Us Lifestyle survey response concerning drinking during the past year. The response options, sourced from AUDIT-C, were collapsed into three study-defined categories: never or monthly or less (code 0); 2–4 times/month or 2–3 times/week (code 1); and ≥ 4 times/week (code 2) [14, 15]. Participants with missing or indeterminate drinking-frequency information were excluded during upstream cohort screening. Code 0 was the reference for both smoking and drinking, and the drinking categories entered models as categorical predictors. The drinking groups describe frequency, not alcohol quantity or AUDIT-C risk scores. Further measurement details are provided in S1 Text. The temporal ordering used throughout the models was

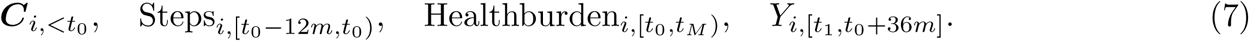

Age and sex at birth were included to account for demographic differences related to physical activity and cardiovascular recording; recorded smoking status and drinking categories represented available pre-baseline health behaviours potentially related to both. Post-baseline health-burden components were not used as substitutes for pre-exposure illness or health-care utilisation.

### Pooled health-burden score

We summarised the device-derived sleep indicator, self-reported mental-health diagnostic history, and EHR clinical burden with one health-burden score constructed using pooled principal component analysis (PCA). The score was a pragmatic composite across three data sources, not a validated latent biological trait. Its interpretation depended on device measurement and coverage, recall and reporting of prior diagnoses, and clinical recording practices. PCA was fitted once in the common *t*_1_-landmark cohort without using the subsequent outcome. Sleep, Mental, and Clinical were separately standardised; the first principal component (PC1) was retained, oriented so that larger values represented greater health burden, and standardised to mean 0 and SD 1. We denote this score by *M*^-^*_i_*. The same pooled score definition was used for all sex and age comparisons. Within each participant-bootstrap sample, domain standardisation and PCA were repeated, PC1 was aligned with the original pooled loading, and the resulting score was standardised again. S1 Fig illustrates the score and fitted models.

The score combines measurements and reports obtained after exposure assessment, but self-reported mental-health diagnoses and EHR clinical conditions may have existed before the exposure window. Separation of ascertainment windows therefore does not establish disease-onset order or show that activity preceded development of these conditions. Unlike latent multiple-mediator methods specifically designed to address unmeasured confounding [16], our PCA was used only for dimension reduction and was not assumed to remove confounding.

### Statistical analysis

Baseline characteristics were summarised as means and standard deviations or counts and percentages, without significance testing. M1 and the exploratory statistical decomposition used the common *t*_1_-landmark cohort and outcome period; analysis-specific subsets and expanded samples are described below. Let ***C****_i_* = {Age*_z,i_,* Sex*_i_,* Smoking*_i_,* Drinking*_i_*} denote the pre-*t*_0_ adjustment set; Steps and age were standardised using the pooled analytic cohort.

#### Primary association and sensitivity analyses

M1 was a three-category multinomial logistic regression with no recorded CVD as the reference category:

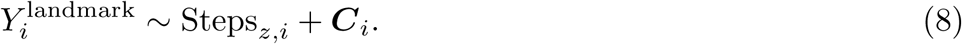

The adjustment variables were selected as available pre-baseline demographic and behavioural covariates; they were not intended to constitute a sufficient adjustment set for causal identification. The acute-care-versus-outpatient estimate was a contrast of coefficients from this same model. A Steps-only multinomial model provided unadjusted estimates using the same participants, outcome categories, and pooled Steps scale (S15 Table). M1 did not adjust for the post-*t*_0_ health-burden score or its components, but its sample remained restricted by the sleep-coverage and survey-completeness requirements. A centred natural cubic spline for Steps was compared with the linear term using a joint likelihood-ratio test (4 df). M1 category probabilities were standardised over the observed covariate distribution across the central 95% of step counts, with simulation-based 95% confidence bands (Fig 2).

**Fig 2.**
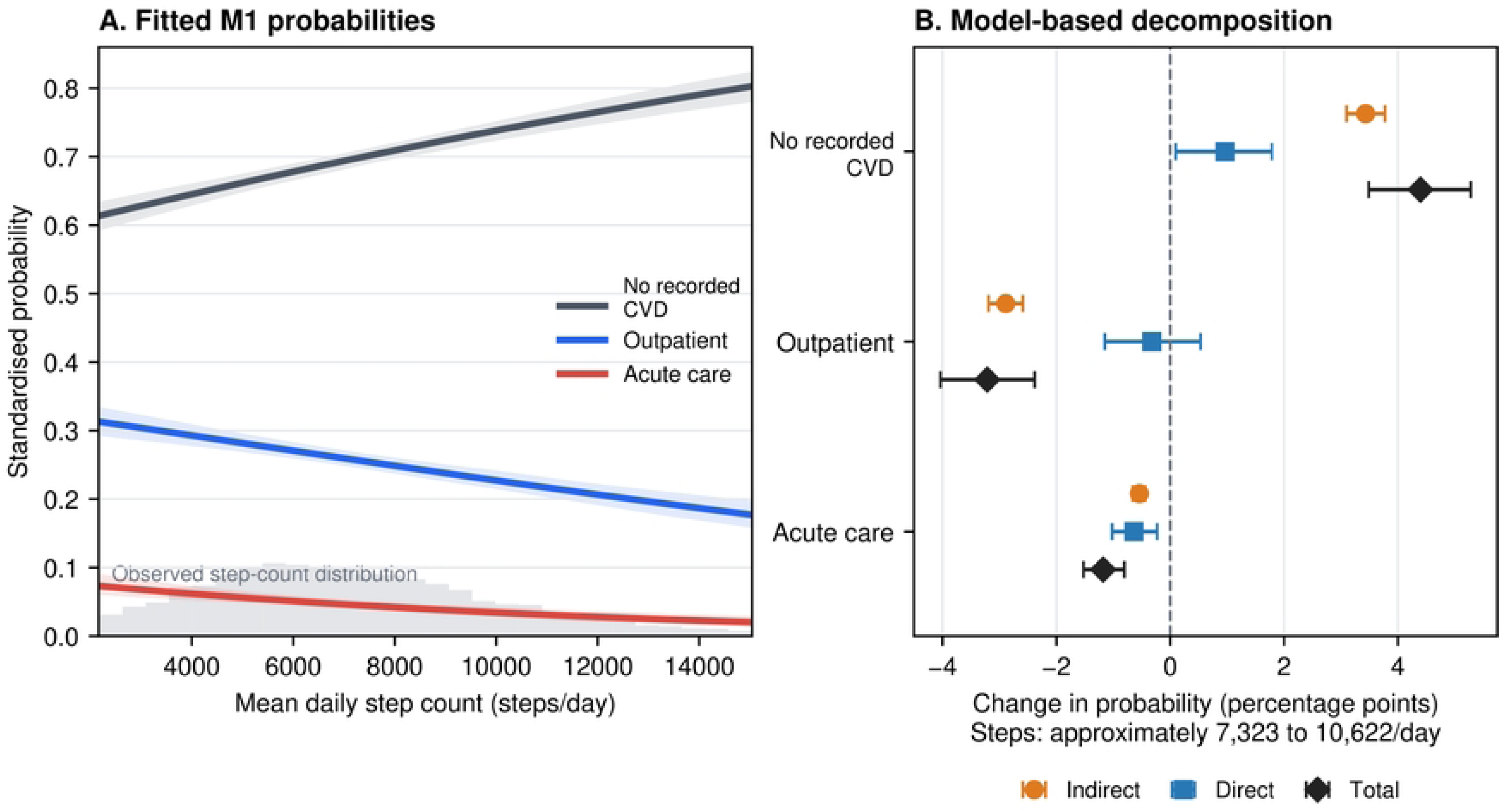
Standardised outcome probabilities and pooled statistical decomposition. Both panels use the common *t*_1_-landmark cohort of 8,292 participants (5,780 participants without recorded CVD, 2,127 outpatient recordings, and 385 acute-care recordings), with outcomes during [*t*_1_*, t*_0_ + 36 months]. (A) Covariate-standardised probabilities from M1 across the central 95% of observed daily step counts. Curves average over the observed age, sex-at-birth, recorded-smoking-status, and drinking-category distribution. Shaded bands are 95% confidence intervals from simulation using the fitted coefficient covariance matrix; grey bars show the step-count distribution. A centred natural-cubic-spline test provided no clear evidence of nonlinearity (4 df; *p* = 0.139), so a linear step-count term was retained on the multinomial-logit scale. (B) Indirect (ACME), direct (ADE), and total probability contrasts obtained by combining predictions from the mediator model and the M2 outcome model, specifically comparing approximately 7323 with 10622 steps/day (the pooled mean with 1 pooled SD above that mean), with 95% participant-bootstrap confidence intervals. Values are percentage-point changes in the original No recorded CVD, Outpatient, and Acute-care probabilities for this exposure contrast, not for every possible 1-SD increase. The three categories share a common denominator and sum to zero within effect type. These are model-based decompositions; pairwise-normalised setting comparisons are reported separately in S3 Table.

Among the 2,512 classified CVD cases, a binary acute-care-versus-outpatient logistic model used the same adjustment set and pooled scaling as M1. This was an associational consistency check under an alternative fitting approach, not independent validation. We described missing-setting and other/unknown cases separately, comparing each subgroup with all classified cases using standardised mean differences (S7 Table, panel A; S1 Text). A separate four-category model retained all 603 unclassified cases (*N* = 8,895); this administrative category did not impute their settings. In a complementary post-hoc analysis, binary logistic models compared any qualifying primary CVD recording, regardless of setting, with no recorded CVD in the original 8,292 participants and in all 8,895 prepared participants. Both used M1’s adjustment set and original Steps and age scales, with Wald 95% confidence intervals. The expanded binary analysis retained all 603 unclassified cases without assigning them a setting (S7 Table).

To examine classification uncertainty directly, a post-hoc analysis added the 455 missing-setting cases (*N* = 8,747), retaining the exclusion of 148 other/unknown cases whose eligibility for either target setting was uncertain. Across 42 scenarios, fractional multinomial models varied the assumed acute proportion and its association with Steps. Intervals from 2000 participant-bootstrap samples were conditional on each allocation scenario, not uncertainty about its validity (Fig 3; S14 Table). These supplementary outcome analyses did not refit the exploratory mediation models.

**Fig 3.**
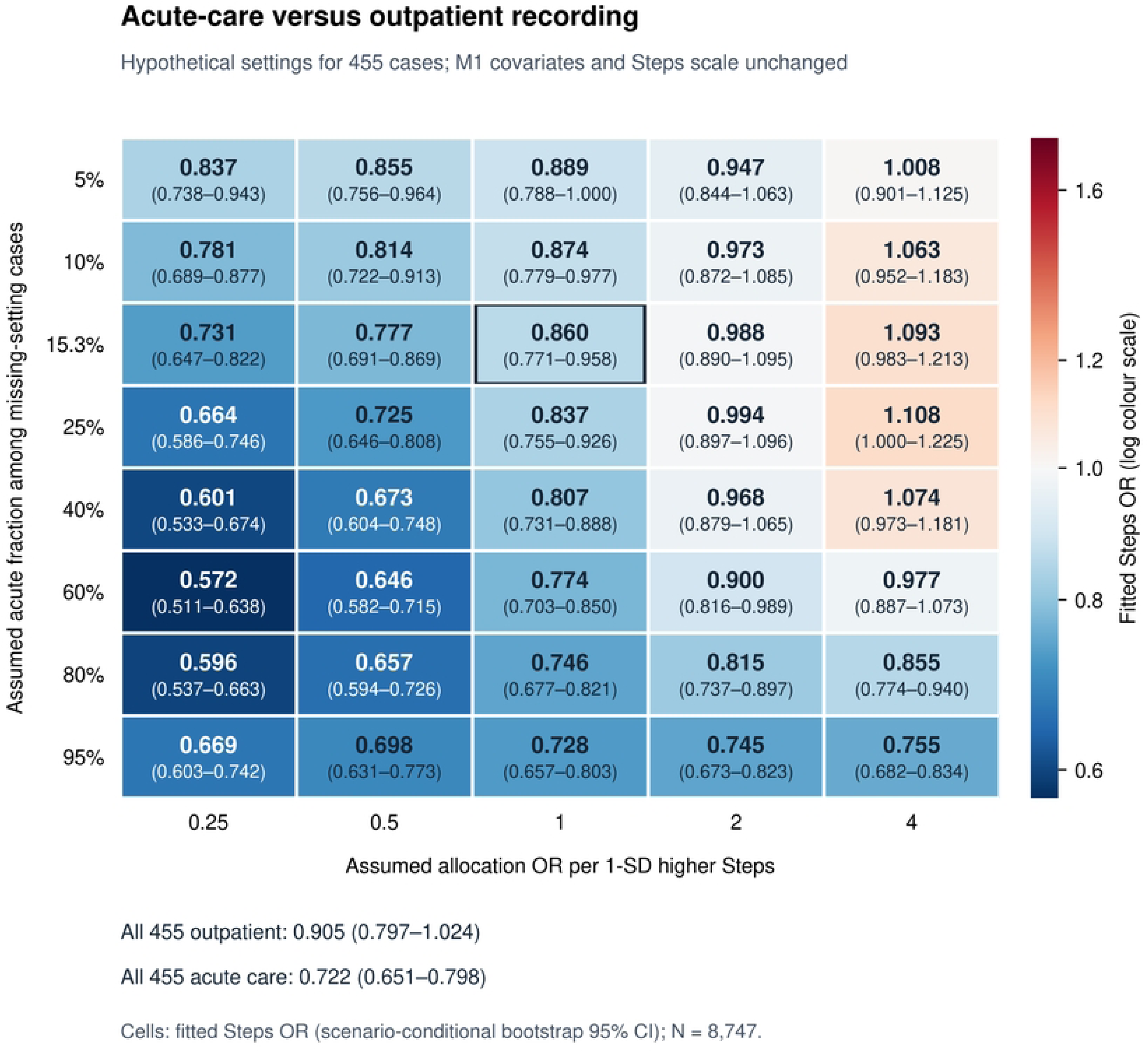
Sensitivity of the Steps association with CVD recording setting to hypothetical allocation of missing settings. Cells report the acute-care-versus-outpatient Steps OR per 1 original pooled SD (3298.82 steps/day), with scenario-conditional 95% confidence intervals from 2000 participant-bootstrap samples. The 455 missing-setting cases were added to the 8,292 classified-cohort participants; 148 other/unknown cases remained excluded. Rows specify the assumed mean acute-care probability among missing-setting cases, and columns specify the assumed acute-allocation OR per 1-SD higher Steps. This allocation OR is distinct from the fitted Steps OR inside each cell. The outlined reference uses the known-setting acute fraction, 385*/*2,512 (15.3%), and Steps-independent allocation (allocation OR=1). Colours represent fitted Steps ORs on a log scale centred at 1, not significance thresholds. The all-outpatient and all-acute scenarios below the grid are not mathematical bounds. All scenarios are hypothetical, and confidence intervals condition on the allocation assumptions rather than their validity. Full methods and all outcome contrasts are in S1 Text and S14 Table. CI=confidence interval; OR=odds ratio.

A post-hoc age-adjustment sensitivity replaced linear age in M1 and the case-only model with a centred natural cubic spline, holding samples, pooled Steps scaling, and other covariates fixed (S8 Table). These analyses did not replace the primary model. Spline specifications, model checks, probability simulations, and the allocation algorithm are detailed in S1 Text.

#### Exploratory statistical decomposition

Using the pooled score *M*^-^*_i_*, we fitted a linear mediator model and a three-category multinomial outcome model (M2):

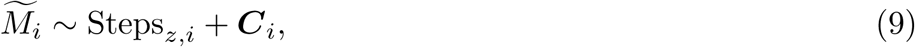

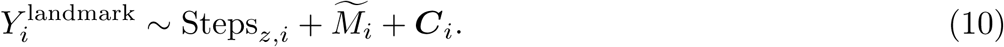

G-computation combined their predictions to estimate indirect (ACME), direct (ADE), and total probability contrasts [17]. The exposure contrast was fixed at Steps*_z_* = 0 versus 1, approximately 7323 versus 10622 steps/day. Predictions were averaged over the observed covariate distribution and integrated over empirical mediator residuals. ACME averaged the indirect contrasts at both exposure reference levels; ADE averaged the direct contrasts under both mediator reference distributions. Their sum equalled the total contrast. Original-category contrasts shared one multinomial denominator and summed to zero across categories within each contrast type; results were reported in percentage points.

For the three pairwise comparisons, probabilities were normalised as *p_j_/*(*p_j_* + *p_k_*) for each participant and residual value before averaging. These contrasts have different denominators, so their magnitudes were not ranked across comparisons. We retain the conventional abbreviations ACME (average causal mediation effect) and ADE (average direct effect) to identify the estimands, but interpret them as exploratory statistical indirect and direct associations. Later ascertainment does not establish disease-onset order, and unmeasured confounding and cohort selection preclude a mechanistic interpretation. M1-to-M2 coefficient changes were not treated as indirect-association estimates because logistic ORs are non-collapsible. Complete definitions and identifying assumptions are provided in S1 Text.

Percentile 95% confidence intervals and two-sided bootstrap tail probabilities used 2000 participant-level resamples. Domain standardisation, score construction, and both regressions were repeated, while original pooled Steps and age scales were fixed. Score-composition sensitivities used equal-weight standardisation, collapsed lipid indicators, and omission of Sleep, with the same participants and paired bootstrap resamples (S4 Table; S10 Table). All score comparisons retained the original sleep-coverage eligibility requirement. Implementation and numerical checks are in S1 Text.

#### Secondary sex and age analyses

M1 sex modification was tested jointly across the two multinomial logits. Exploratory pooled moderated-mediation models evaluated sex and continuous-age interactions for the Steps–score, residual Steps–outcome, and score–outcome associations [18]. Female-minus-Male and age-64-minus-age-50 ACME differences were standardised to the same pooled covariate population; a Sex-by-Age difference-in-differences analysis was exploratory. Flexible-age moderation and independently fitted sex-stratified models were supplementary, retaining a common pooled score definition. Model specifications and bootstrap procedures are in S1 Text. Coefficient interactions and probability-scale differences test distinct hypotheses in nonlinear models; subgroup significance alone was not evidence of heterogeneity.

All tests were two-sided. Holm adjustment was applied separately to three families: (i) six pathway-interaction tests, sex and linear age for each of the three associations above (S5 Table); (ii) six ACME-difference tests, Female minus Male and age 64 minus age 50 for each of the three outcome comparisons (S6 Table); and (iii) three exploratory Sex-by-Age ACME difference-in-differences tests, one per comparison. Each outcome-path test jointly assessed both nonreference logits and counted once. The separate M1 global Steps-by-sex test and S5’s flexible-age and highest-order Sex-by-Age pathway tests were outside these families and reported unadjusted. Bootstrap intervals were pointwise, not multiplicity-adjusted simultaneous intervals. Analyses used Python with pandas, NumPy, SciPy, statsmodels, and patsy; package versions are reported in S1 Text.

## Data and code availability

The study used Registered Tier Curated Data Repository version 8 of the *All of Us* Research Program. Participant-level data cannot be redistributed by the authors under the programme’s data-use requirements. Eligible researchers can apply independently through https://www.researchallofus.org/register/.

Access requires an institutional Data Use and Registration Agreement covering the Registered Tier, researcher registration and identity verification, the required training, and agreement to the Data User Code of Conduct. Approved researchers access and analyse these data within the secure Researcher Workbench; author-mediated transfer of participant-level data is not an access route.

A machine-readable workbook of the reported supplementary table results is provided in S1 Dataset. It contains aggregate estimates and summaries at the precision shown in S1–S15 Tables, not participant-level data.

Downstream analysis code, variable definitions, and environment documentation are publicly available at https://github.com/kaixiang164-byte/fitbit-cvd-analysis (commit 9a3bfb8). The release includes analysis scripts, reporting utilities, synthetic tests, and a vocabulary-only Clinical concept exporter. The analysis code starts from a prepared analytic cohort and does not implement upstream participant extraction or eligibility filtering. Participant-level data are not included and require independent authorised access to the All of Us Researcher Workbench as described above.

## Results

### Study population and landmark-aligned characteristics

The reported starting cohort included 10,365 participants without primary or secondary CVD recorded in their available pre-*t*_0_ EHR history, after applying the diagnosis-status, demographic-completeness, sleep-coverage, survey-completeness, recorded-smoking-status, and drinking-frequency eligibility requirements. Of these, 1,470 with primary or secondary CVD recorded during [*t*_0_*, t*_1_) were excluded upstream, leaving 8,895 retained participants: 5,780 participants without recorded CVD with neither diagnosis status recorded through *t*_0_ + 36 months and 3,115 with a first qualifying primary CVD record on or after *t*_1_. Among these cases, 2,127 had outpatient recording and 385 had acute-care recording; 148 had another or unknown setting and 455 had missing setting. Excluding the latter 603 yielded the common association and exploratory mediation cohort of 8,292 participants (5,780 participants without recorded CVD, 2,127 outpatient, and 385 acute-care recordings; Fig 1). The no-recorded-CVD category denotes absence of primary or secondary records, not confirmed absence of disease; endpoint EHR contact does not establish continuous capture.

Participant characteristics are organised by their position in the landmark timeline in Table 1: age, sex, recorded smoking status, drinking categories, and Steps precede *t*_0_, whereas device-derived sleep, self-reported mental-health diagnostic history, and EHR clinical burden were ascertained during the one-month [*t*_0_*, t_M_*) health-burden ascertainment window. Mean age was 57.0 years (SD 14.3) among participants without recorded CVD, 67.1 years (SD 12.9) among participants with an outpatient recording, and 63.5 years (SD 14.4) among those with an acute-care recording. Corresponding mean daily step counts were 7548.8 (SD 3305.2), 6868.3 (SD 3228.5), and 6451.1 (SD 3188.5), respectively.

**Table 1.**
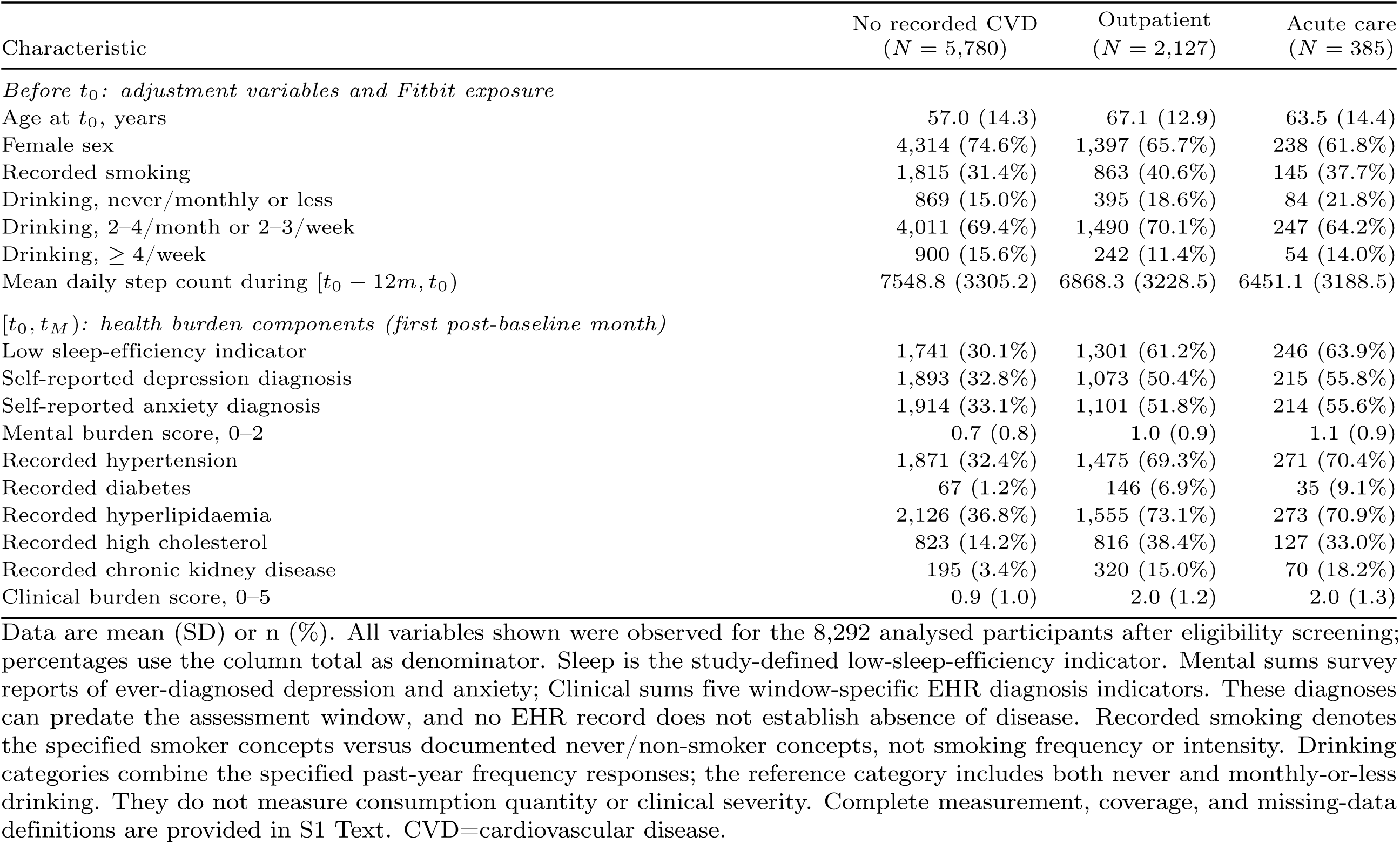
Participant characteristics by cardiovascular recording and landmark measurement window.

### Association between daily steps and cardiovascular recording

The M1 primary multinomial analysis included the common *t*_1_-landmark cohort of 8,292 participants and adjusted only for age, sex, recorded smoking status, and the defined drinking categories; it did not include the post-exposure health-burden score or any of its components. Each 1-SD higher mean daily step count (3298.82 steps/day) was associated with lower odds of outpatient recording versus no recorded CVD (OR 0.786, 95% CI 0.744–0.830; *p <* 0.001), acute-care recording versus no recorded CVD (OR 0.659, 95% CI 0.586–0.742; *p <* 0.001), and acute-care versus outpatient recording (OR 0.839, 95% CI 0.742–0.948; *p* = 0.005; Table 2). These estimates describe adjusted associations among participants without recorded CVD before *t*_1_. Unadjusted estimates from the same cohort were also inverse for all three comparisons; these are presented alongside M1 in S15 Table.

**Table 2.**
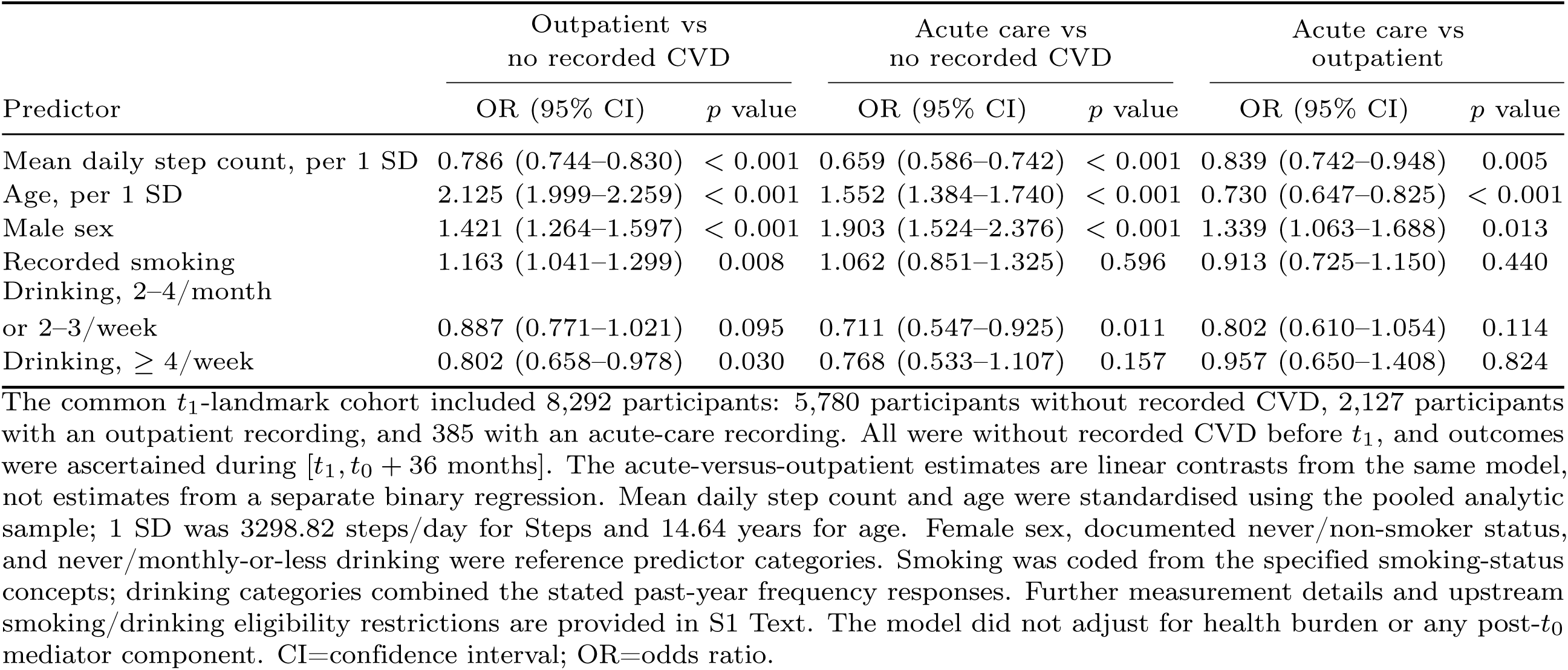
M1 primary multinomial logistic regression of cardiovascular recording category.

Alternative fitting approaches and age specifications yielded similar Steps associations. The case-only acute-care-versus-outpatient estimate with linear age adjustment (OR 0.851, 95% CI 0.755–0.960; *p* = 0.009) was concordant with the corresponding M1 estimate (OR 0.839). Natural cubic spline adjustment for age improved overall M1 fit but left the Steps estimates similar across all three contrasts; the acute-care-versus-outpatient OR was 0.841 (0.743–0.951). These analyses provide consistency checks under alternative model specifications (S8 Table). Separately, the Steps spline test provided no clear evidence of nonlinearity (*p* = 0.139), so the graphical summary retained a linear Steps term on the multinomial-logit scale.

### Sensitivity to incomplete recording settings

Among the 3,115 qualifying CVD cases, 603 (19.4%) had unclassified settings: 455 missing and 148 other/unknown. Missing-setting cases had lower mean daily step counts than the 2,512 classified cases (5619.1 versus 6804.4 steps/day; standardised mean difference −0.365), whereas other/unknown cases had a similar mean (6778.1 steps/day; standardised mean difference −0.008). S7 Table, panel A reports all four case groups separately, including step-count medians and quartiles. Retaining all 603 unclassified cases as a fourth outcome category left the known-setting associations similar, including acute-care versus outpatient recording (OR 0.841, 95% CI 0.745–0.950; *p* = 0.005). The step-count OR for the combined unclassified category versus no recorded CVD was 0.556 (95% CI 0.502–0.616; S7 Table, panel B). This supports stability under the expanded model, not robustness to unknown acute-care or outpatient assignments. The descriptive differences indicate that missing-setting cases are not interchangeable with classified cases on measured characteristics; they do not identify the missingness mechanism or quantify selection bias.

When CVD recordings were combined irrespective of setting, the adjusted Steps OR per original pooled SD was 0.765 (95% CI 0.726–0.807) in the 8,292-person cohort (2,512 cases) and 0.727 (0.692–0.764) in the 8,895-person prepared cohort (3,115 cases); both included the same 5,780 participants without recorded CVD (S7 Table, panel C). Thus, the inverse association with this combined recording endpoint persisted when all unclassified cases were retained without setting allocation. These nested-cohort estimates do not resolve uncertainty in the acute-care-versus-outpatient comparison.

In the allocation sensitivity, assigning an expected 15.3% of the 455 missing-setting cases to acute care independently of Steps yielded an acute-care-versus-outpatient OR of 0.860 (scenario-conditional 95% bootstrap CI 0.771–0.958). This proportion matched the known-setting cases only as a fixed reference. Holding it fixed but doubling acute-assignment odds per 1-SD higher Steps attenuated the OR to 0.988 (0.890–1.095); quadrupling these odds shifted the point estimate above one (1.093, 0.983–1.213), without establishing a positive association. Thus, the magnitude of the association moved toward the null or changed direction, rather than merely losing statistical significance (Fig 3). Both associations for either CVD recording setting versus no recorded CVD remained inverse across the tested scenarios (S14 Table). The 148 other/unknown cases remained excluded. These scenarios tested a limited class of allocation assumptions, allowing allocation to depend on Steps but not on age, comorbidity, or health-care access; they do not establish robustness to all plausible missingness mechanisms.

### Exploratory pooled indirect-association results

The pooled health-burden score captured 51.26% of the standardised domain variance; each 1-SD higher daily step count was associated with a 0.228-SD lower score (coefficient −0.228, 95% CI −0.249 to −0.208; *p <* 0.001). Comparing approximately 7323 with 10622 steps/day, the model-based indirect probability contrasts were 3.433 (3.097 to 3.772) percentage points for No recorded CVD, −2.890 (−3.192 to −2.589) for Outpatient, and −0.543 (−0.659 to −0.438) for Acute care. Table 3 and Fig 2 report the original-category decomposition; score loadings and M2 coefficients are in S12 Table and S13 Table.

**Table 3.**
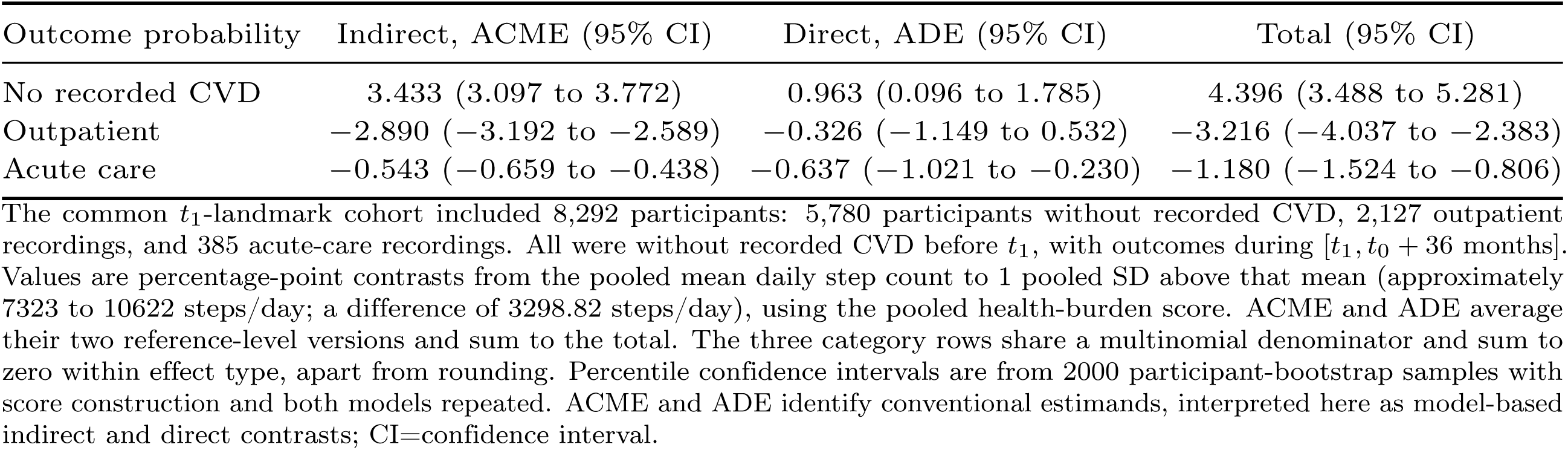
Pooled statistical decomposition on the original three-category probability scale.

| Outcome probability | Indirect, ACME (95% CI) | Direct, ADE (95% CI) | Total (95% CI) |
| --- | --- | --- | --- |
| No recorded CVD | 3.433 (3.097 to 3.772) | 0.963 (0.096 to 1.785) | 4.396 (3.488 to 5.281) |
| Outpatient | $-2.890$ ($-3.192$ to $-2.589$) | $-0.326$ ($-1.149$ to $0.532$) | $-3.216$ ($-4.037$ to $-2.383$) |
| Acute care | $-0.543$ ($-0.659$ to $-0.438$) | $-0.637$ ($-1.021$ to $-0.230$) | $-1.180$ ($-1.524$ to $-0.806$) |

The common $t_1$ -landmark cohort included 8,292 participants: 5,780 participants without recorded CVD, 2,127 outpatient recordings, and 385 acute-care recordings. All were without recorded CVD before $t_1$ , with outcomes during $[t_1, t_0 + 36 \text{ months}]$ . Values are percentage-point contrasts from the pooled mean daily step count to 1 pooled SD above that mean (approximately 7323 to 10622 steps/day; a difference of 3298.82 steps/day), using the pooled health-burden score. ACME and ADE average their two reference-level versions and sum to the total. The three category rows share a multinomial denominator and sum to zero within effect type, apart from rounding. Percentile confidence intervals are from 2000 participant-bootstrap samples with score construction and both models repeated. ACME and ADE identify conventional estimands, interpreted here as model-based indirect and direct contrasts; CI=confidence interval.

Pairwise-normalised indirect contrasts were −3.223 (−3.544 to −2.904) percentage points for outpatient versus no recorded CVD and −1.341 (−1.571 to −1.121) for acute care versus no recorded CVD. The acute-care-versus-outpatient indirect contrast was inconclusive (−0.185 (−0.496 to 0.153) percentage points; *p* = 0.315), although its direct and total contrasts were inverse (S3 Table). These pairwise contrasts have different denominators and were not ranked by magnitude.

Equal weighting and combining the two lipid indicators yielded similar indirect-contrast patterns (S4 Table). Omitting Sleep changed the pairwise-normalised indirect contrast for outpatient versus no recorded CVD from −3.223 to −3.041 percentage points and that for acute care versus no recorded CVD from −1.341 to −1.257. Paired changes were 0.182 (95% CI −0.001 to 0.364) and 0.084 (−0.025 to 0.193) percentage points, respectively, for the same fixed Steps contrast. With a score constructed from Mental and Clinical alone, both indirect contrasts remained inverse, while the acute-care-versus-outpatient indirect contrast remained inconclusive (S10 Table).

### Secondary sex and age analyses

The M1 global Steps-by-sex test provided no clear evidence of interaction (*p* = 0.301; S11 Table), and none of the six sex and linear-age pathway-interaction tests met the Holm-adjusted threshold (smallest adjusted *p* = 0.053; S5 Table). Probability-scale differences in indirect contrasts meeting the Holm-adjusted threshold occurred only in comparisons of either CVD recording setting with no recorded CVD: the Female-minus-Male acute-versus-no-recorded-CVD contrast indicated a more inverse estimate among males, and age-related differences occurred in comparisons of both CVD recording settings with no recorded CVD. For acute versus outpatient recording, neither the sex nor the age comparison met this threshold (S6 Table); the age grid and independently fitted sex-specific estimates are supplementary (S1 Table and S2 Table). In a nonlinear outcome model, probability-scale contrasts depend on fitted outcome probabilities as well as pathway coefficients, so they can differ even when coefficient-interaction tests do not meet significance thresholds.

These tests address different null hypotheses and do not establish sex- or age-specific biological mechanisms.

## Discussion

Higher daily step counts were associated with lower odds of subsequent CVD recording in both outpatient and acute-care settings relative to no recorded CVD after adjustment for measured covariates. Both associations remained inverse across the tested missing-setting allocations, whereas the stronger acute-care association among classified cases depended on allocation assumptions. Exploratory model-based analyses identified indirect associations involving the health-burden score in comparisons with participants without recorded CVD, but evidence for the between-setting indirect contrast was inconclusive.

Previous studies have associated objectively measured activity with cardiovascular outcomes [1–3], including Fitbit-linked chronic disease research in All of Us [4]. This study extends aggregate CVD comparisons by distinguishing outpatient and acute-care recording and quantifying how the resulting associations vary under assumptions about missing settings. Combining all qualifying primary CVD recordings also yielded an inverse Steps association without assigning the unclassified cases to a setting. The distinction matters for wearable–EHR research: an association with CVD recording does not by itself establish a stable difference between recording settings. The allocation analysis describes this dependence without identifying the true missingness mechanism. These endpoints concern clinical documentation, not disease onset or validated severity.

The pooled health-burden score summarised clinically related domains [5–8], with PC1 explaining 51.26% of the standardised variance. Similar indirect-association patterns under equal weighting, combined lipid indicators, and omission of Sleep support stability under these scoring choices, not a validated biological mechanism. Self-reported diagnostic history and EHR conditions may predate exposure, while device measurements, recall, and clinical recording capture different aspects of health. These domains differ in both their clinical meaning and measurement processes. PCA cannot distinguish shared health variation from source-specific recording processes; alternative integration methods would also require measurement validation [19].

The secondary analyses provided no clear evidence of sex or age differences in the acute-care-versus-outpatient indirect contrast. Probability-scale differences in comparisons of CVD recording settings with no recorded CVD do not establish demographic-specific biological pathways; menopausal status was not measured, precluding menopause-specific inference.

A strength of this study is the integration of wearable activity measured over a prespecified year with subsequent EHR recording setting. The primary association and exploratory mediation models shared the cohort, exposure scale, and outcome interval, with one pooled score definition. Original-category probability summaries provide a common denominator, while pairwise contrasts directly compare recording settings. Future studies should combine repeated activity measurements with more complete phenotype, recording-setting, and pre-exposure health information.

Several limitations remain. First, differences in CVD subtype composition could contribute to the acute-care-versus-outpatient comparison. The broad endpoint included heart failure, arrhythmias, cerebrovascular disease, venous thromboembolism, and structural disorders, rather than being restricted to atherosclerotic CVD or major adverse cardiovascular events. Outcome classification used the first qualifying primary-coded CVD record, which did not establish biological onset or equivalent disease severity across settings. Participant-level subtype and the expanded historical descendant code list were not retained, limiting independent verification. S9 Table lists the defining disease concepts, descendant inclusion, and diagnosis-status codes used in screening. No same-day multiple-setting qualifying primary CVD diagnoses were identified upstream, although the prepared dataset did not retain all source encounters for re-audit. Outpatient recording required one qualifying code, not repeat-code confirmation.

Second, adjustment was limited to age, sex at birth, recorded smoking status, and the defined drinking categories. Race and ethnicity were unavailable, leaving potential residual confounding related to associated social and structural factors and preventing assessment of cohort representation and generalisability across racial and ethnic groups. The binary smoking indicator combined smoking concepts of differing intensity and did not quantify duration or cumulative exposure; the drinking categories did not capture alcohol quantity or episodic heavy drinking, leaving potential residual confounding. Comorbidity and health-care utilisation measures from before the step-count exposure window were unavailable in the analytic extract, precluding a sensitivity analysis additionally adjusting for these factors. The subsequently measured health-burden score does not substitute for these covariates. Prior illness, health-care utilisation, and socioeconomic circumstances may account for the observed associations. Pre-existing illness that both reduces activity and increases CVD recording could exaggerate an inverse association, whereas differential access and cohort selection could bias estimates in either direction; these contributions could not be quantified. Health-care access and site could also confound the associations; calendar and seasonal variation were not evaluated. Future studies should prioritise dated diagnoses and encounter histories preceding the start of the Steps exposure window (*t*_0_ − 12 months), characterise EHR coverage, and evaluate additional adjustment for prior illness and health-care utilisation.

Third, all analyses, including M1, were restricted to participants without recorded CVD before *t*_1_ who also met post-baseline sleep-coverage and survey-completeness requirements and the pre-baseline smoking-status and drinking-frequency eligibility criteria. Excluding explicit former smokers and participants without classifiable smoking or drinking information further restricted the study population. The reported flow begins after eligibility screening; counts for earlier demographic-completeness, sleep-coverage, survey-completeness, smoking-status, drinking-frequency, and diagnosis-status exclusions, including their ordering and overlap, were not retained. Variable-specific missingness counts before eligibility screening were also unavailable, so completeness in the analysed cohort does not describe missingness before screening. Selection from a broader eligible population could not be quantified. The 1,470 upstream exclusions define the landmark population but could induce selection related to activity or health status. Conditioning on recorded case status can also introduce selection or collider bias in case-only comparisons. EHR ascertainment depends on health-care contact and recording completeness: the no-recorded-CVD category denotes absence of primary or secondary records, not verified absence of disease. Requiring an EHR record on or after the final endpoint selected participants with continuing contact but did not establish continuous capture of events during follow-up. No deaths were recorded in the analytic dataset, but completeness of mortality ascertainment could not be established.

Fourth, missing-setting cases had lower mean step counts than classified cases, whereas other/unknown cases had similar mean step counts. These descriptive differences do not identify why settings were unclassified. The allocation scenarios demonstrated sensitivity of the setting comparison but were not estimates of the actual missingness mechanism: they assumed that all 455 missing-setting cases belonged to one of the two target settings, excluded the 148 other/unknown cases, and allowed allocation to vary only with Steps. Confidence intervals were conditional on fixed scenarios and did not capture uncertainty about their validity. The true setting distribution and resulting selection bias therefore remain unresolved.

Fifth, Fitbit users are a selected population, and requiring at least nine valid Steps months does not eliminate intermittent recording or device-related measurement error [20]. Sleep coverage during the health-burden ascertainment month is distinct from Steps coverage before baseline. At least seven valid main-sleep days were required, but this does not establish complete monthly coverage. The sleep-efficiency threshold was study-specific and not clinically validated. The post-baseline event-free year provides a lag, but cannot eliminate reverse causation from preclinical disease. Finally, model specification and unmeasured confounding limit the interpretation of the indirect associations; only 385 participants had acute-care recording, limiting precision for secondary comparisons.

In conclusion, after adjustment for the measured covariates, daily step count was inversely associated with both CVD recording settings relative to no recorded CVD. These associations retained their direction across the tested missing-setting allocations, but the stronger acute-care association in the classified cohort was sensitive to allocation assumptions. Exploratory analyses identified statistical indirect associations involving measured health burden in comparisons with participants without recorded CVD, while the acute-versus-outpatient indirect contrast remained inconclusive. Together, these findings highlight the role of endpoint definition and recording completeness in interpreting wearable–EHR associations. Replication with more complete setting, phenotype, and observation information is needed before clinical application.

## Acknowledgments

We gratefully acknowledge All of Us participants for their contributions, without whom this research would not have been possible. We also thank the National Institutes of Health’s All of Us Research Program for making available the participant data examined in this study.

## Supporting information

**S1 Text. Supplementary measurement definitions and statistical methods.** Cohort-construction rules; complete sleep, survey, clinical-burden, and behavioural-covariate definitions; a consolidated table of data sources, variable derivation, and analysis-code coverage; spline and probability-curve specifications; probability-decomposition formulas; secondary sex/age models and multiplicity; bootstrap implementation and software versions; and the missing-setting allocation algorithm and conditional uncertainty.

**S1 Fig. Construction and analysis of the pooled PCA-derived health-burden score.** Panel A shows separate standardisation of device-derived Sleep, self-reported Mental diagnostic history, and EHR Clinical burden ascertained during [*t*_0_*, t_M_*), followed by one pooled PCA. Complete measurement, coverage, and missing-data definitions are provided in S1 Text. Panel B shows the mediator and multinomial outcome-model structure with pre-*t*_0_ adjustment. The common *t*_1_-landmark cohort included 8,292 participants: 5,780 participants without recorded CVD, 2,127 outpatient recordings, and 385 acute-care recordings. CVD=cardiovascular disease; EHR=electronic health record; PCA=principal component analysis; PC1=first principal component.

**S1 Table. Conditional model-based indirect contrasts across the five-age evaluation grid.** Values are conditional ACME estimates with 95% bootstrap confidence intervals, expressed as pairwise-normalised percentage-point differences comparing the pooled mean daily Steps with 1 pooled SD above that mean (approximately 7323 to 10622 steps/day), not an arbitrary 1-SD increase. The common *t*_1_-landmark cohort included 8,292 participants: 5,780 participants without recorded CVD, 2,127 outpatient recordings, and 385 acute-care recordings. These are model-based evaluations from the continuous linear-age moderation model, standardised over the same pooled covariate distribution, rather than estimates from independently fitted age strata. Each indirect contrast for either CVD setting versus no recorded CVD had *p* = 0.001; all acute-versus-outpatient confidence intervals included zero.

**S2 Table. Independently fitted sex-stratified model-based mediation contrasts.** Values are sex-specific model-based indirect (ACME), direct (ADE), and total contrasts with 95% bootstrap confidence intervals, expressed as pairwise-normalised percentage-point differences comparing the pooled mean daily Steps with 1 pooled SD above that mean (approximately 7323 to 10622 steps/day), not sex-specific starting levels. The female analysis included 5,949 participants (4,314 participants without recorded CVD, 1,397 outpatient recordings, and 238 acute-care recordings); the male analysis included 2,343 participants (1,466 participants without recorded CVD, 730 outpatient recordings, and 147 acute-care recordings). One pooled PCA definition and the original pooled Steps and age scales were retained, but the mediator and multinomial outcome models were fitted independently within females and males. These estimates are descriptive and supplementary; formal sex heterogeneity was evaluated with the exploratory pooled moderated-mediation models.

**S3 Table. Complementary pairwise-normalised model-based mediation contrasts.** ACME, ADE, and total percentage-point contrasts with 95% participant-bootstrap confidence intervals compare outpatient versus no recorded CVD, acute versus no recorded CVD, and acute versus outpatient recording. The common *t*_1_-landmark cohort included 8,292 participants: 5,780 participants without recorded CVD, 2,127 outpatient recordings, and 385 acute-care recordings. The exposure contrast is from the pooled mean daily step count to 1 pooled SD above it. Normalisation is performed before covariate and residual averaging; denominators differ across comparisons, so effect magnitudes are not ranked across rows.

**S4 Table. Sensitivity of model-based indirect contrasts to health-burden-score construction.** Pairwise-normalised ACMEs are compared across the primary pooled PCA score, an equal-weight standardised composite, and a pooled PCA score using a Clinical burden with hyperlipidaemia and high cholesterol collapsed to one indicator. All analyses used the common *t*_1_-landmark cohort of 8,292 participants: 5,780 participants without recorded CVD, 2,127 outpatient recordings, and 385 acute-care recordings.

**S5 Table. Formal sex and age interaction tests in the exploratory mediation models.** All models used the common *t*_1_-landmark cohort. One Holm family comprised six tests: sex and linear age each modifying the Steps–score, residual Steps–outcome, and score–outcome associations. Each outcome-path test was joint across the two nonreference logits. Flexible-age and highest-order Sex-by-Age pathway tests were sensitivity or exploratory analyses with unadjusted *p* values. These parameter-scale tests differ from the probability-scale ACME comparisons in S6 Table.

**S6 Table. Conditional model-based indirect contrasts and moderator differences.** Values are pairwise-normalised percentage-point differences for the fixed pooled-mean to pooled-mean-plus-1-SD Steps contrast (approximately 7323 to 10622 steps/day), standardised over the same pooled covariate population at the compared moderator levels. The six Female-minus-Male and age-64-minus-age-50 ACME-difference tests across three outcome comparisons formed one Holm family; the three exploratory Sex-by-Age ACME difference-in-differences tests formed another. The 95% intervals are pointwise bootstrap percentile intervals; Holm adjustment applies to the *p* values, not the intervals. Probability-scale contrasts can vary even when parameter-scale interaction tests do not meet significance thresholds and do not establish biological mechanisms.

**S7 Table. Unclassified-setting sensitivity and combined CVD-recording analyses.** Panel A separately describes 2,127 outpatient, 385 acute-care, 455 missing-setting and 148 other/unknown cases, including step-count means, SDs, medians and quartiles. Standardised mean differences compare each unclassified subgroup with all 2,512 classified cases. Small counts and complementary cells are suppressed to prevent disclosure. Panel B compares step-count odds ratios from the original three-category M1 model (*N* = 8,292) and an expanded four-category model (*N* = 8,895). Panel C reports supplementary binary logistic estimates for any qualifying primary CVD recording, regardless of setting, versus no recorded CVD in these same two populations: 2,512 and 3,115 cases, respectively, with the same 5,780 participants without recorded CVD. All models retain M1’s original pooled scaling and adjustment set; no participants were additionally excluded for missing covariates. Panel C does not allocate missing settings, and the nested samples are not independent replications. Neither the fourth-category nor the combined-endpoint analysis resolves the unknown acute-care or outpatient assignments; no exploratory mediation model was re-fitted.

**S8 Table. Sensitivity of Steps associations to nonlinear age adjustment.** Panel A compares Steps ORs and 95% CIs under linear and natural-cubic-spline age adjustment in M1 (*N* = 8,292: 5,780 participants without recorded CVD, 2,127 outpatient recordings, and 385 acute-care recordings) and the case-only model (*N* = 2,512: 2,127 outpatient and 385 acute-care recordings). Panel B reports within-model likelihood-ratio tests and AIC values. Samples, pooled Steps scaling, and other covariates were unchanged. This post-hoc sensitivity did not replace the primary M1 or refit the exploratory mediation models.

**S9 Table. CVD phenotype concepts and diagnosis-status criteria.** Panel A lists 13 All of Us disease concept IDs defining 12 overlapping disease families; the phenotype included these concepts and their descendants. Descendant codes are not individually enumerated. Panel B lists the condition_status_concept_id values used upstream: 32902 (Primary diagnosis) and 32908 (Secondary diagnosis). Both were used for participant screening and eligibility for the no-recorded-CVD category; only primary records determined case timing and setting. Participants with secondary CVD preceding their first primary CVD record were excluded. Participants without recorded CVD had neither primary nor secondary CVD through the endpoint. Full eligibility rules are described in the Methods.

**S10 Table. Sensitivity of pooled model-based mediation contrasts to omitting the Sleep domain.** The primary three-domain pooled PCA score is compared with a pooled PCA score using Mental and Clinical only in the same 8,292 participants (5,780 participants without recorded CVD, 2,127 outpatient recordings, and 385 acute-care recordings). Model-based probability contrasts compare approximately 7323 with 10622 steps/day. The analyses use the same 2000 participant-bootstrap resamples; paired differences compare the two-domain with the three-domain ACME. This sensitivity addresses score composition within the selected cohort, not measurement validity, a sleep-specific indirect effect, or selection due to sleep coverage.

**S11 Table. Sex-specific Steps associations and formal Steps-by-sex interaction contrasts.** Conditional Steps odds ratios for females and males and male-to-female ratios of odds ratios are derived from one pooled multinomial model in the common *t*_1_-landmark cohort of 8,292 participants (5,780 participants without recorded CVD, 2,127 outpatient recordings, and 385 acute-care recordings). The model includes Steps-by-sex interactions and adjusts for age, recorded smoking status, and drinking. The joint two-parameter interaction test is reported separately from subgroup-specific slopes.

**S12 Table. Pooled principal-component characteristics of the health-burden score.** Domain means, standard deviations, and PC1 loadings are reported for the common *t*_1_-landmark cohort of 8,292 participants (5,780 participants without recorded CVD, 2,127 outpatient recordings, and 385 acute-care recordings). Sleep, Mental, and Clinical burden were separately standardised before one pooled PCA; PC1 explained 51.26% of variance and was oriented so that higher scores represented greater health burden.

**S13 Table. M2 multinomial outcome-model associations with Steps and health burden.** Adjusted odds ratios and 95% confidence intervals are shown for Steps and health burden in one multinomial model fitted in the common *t*_1_-landmark cohort of 8,292 participants (5,780 participants without recorded CVD, 2,127 outpatient recordings, and 385 acute-care recordings). The model adjusts for age, sex, recorded smoking status, and drinking; outcomes occur during [*t*_1_*, t*_0_ + 36 months]. Acute-care-versus-outpatient estimates are contrasts from the same model. Coefficient differences from M1 are not interpreted as indirect-association estimates.

**S14 Table. Steps associations under hypothetical missing-setting allocations.** All 42 scenarios report the three M1 outcome contrasts per original pooled SD of Steps (3298.82 steps/day), with scenario-conditional 95% percentile intervals from 2000 participant bootstrap samples. The analysis includes 8,747 participants and excludes 148 other/unknown cases. M1 covariates and standardisation are retained; PCA and the exploratory mediation models are unchanged. Intervals do not quantify uncertainty about allocation assumptions.

**S15 Table. Unadjusted and adjusted step-count associations with cardiovascular recording.** Steps-only and M1-adjusted multinomial estimates are reported in the same 8,292 participants (5,780 participants without recorded CVD, 2,127 outpatient recordings, and 385 acute-care recordings), per original pooled SD of Steps (3298.82 steps/day). M1 adjusts for age, sex at birth, recorded smoking status, and drinking frequency. Acute-care-versus-outpatient estimates use the full joint coefficient covariance. Two-sided Wald 95% confidence intervals and *p* values are shown; changes between models are not interpreted as the proportion explained by confounding.

**S1 Dataset. Machine-readable supplementary table results.** An Excel workbook containing the aggregate results and concept definitions in S1–S15 Tables, with an index and one worksheet per table. Worksheets include table identifiers, units, interval bounds, reported precision, and source descriptions. The workbook contains no participant-level records, individual predictions, or resampling indices.

**S1 Checklist. STROBE checklist for this observational cohort study.**

